# Survival and predictors of deaths of patients hospitalized due to COVID-19 from a retrospective and multicenter cohort study in Brazil

**DOI:** 10.1101/2020.06.07.20125047

**Authors:** Marquiony M Santos, Eudes ES Lucena, Kenio C Lima, Andiara AC Brito, Monica B Bay, Diego Bonfada

## Abstract

The epidemic caused by COVID-19 in Brazil is associated with an unfavorable political scenario, aggravated by intense social inequality and low number of available hospital beds. Therefore, this study aimed to analyze the survival of patients admitted to Brazilian hospitals due to the COVID-19 and estimate prognostic factors. This is a retrospective, multicenter cohort study, based on data from 46285 hospitalizations for COVID-19 in Brazil. Survival functions were calculated using the Kaplan-Meier’s method. The Log-rank test compared the survival functions for each variable and from that, hazard ratios were calculated and the proportional hazards model was used in Cox multiple regression. The smallest survival curves were the ones for patients at the age of 68 years or more, black / brown race, illiterate, living in the countryside, dyspnea, respiratory distress, influenza-like outbreak, O_2_ saturation <95%, X-ray change, length of stay in the ICU, invasive ventilatory support, previous heart disease, pneumopathy, diabetes, down’s syndrome, neurological disease and kidney disease. Better survival was observed in the symptoms and in an asthmatic patient. The multiple model for increased risk of death when they were admitted to the ICU HR 1.28 (95% CI 1.21–1.35), diabetes HR 1.17 (95% CI 1.11–1.24), neurological disease HR 1.34 (95% CI 1.22–1.46), kidney disease HR 1.11 (95% CI 1.02–1.21), heart disease HR 1.14 (95% CI 1.08–1.20), black or brown race of HR 1.50 (95% CI 1.43–1.58), asthma HR 0.71 (95% CI 0.61–0.81) and pneumopathy HR 1.12 (95% CI 1.02–1.23). The overall survival time was low in hospitalizations for COVID-19 and this reinforces the importance of sociodemographic and clinical factors as a prognosis for death. The lack of a protocol for scientific clinical management puts a greater risk of death for about 80 million Brazilians, who are chronically ill or living in poverty. COVID-19 can promote selective mortality that borders the eugenics of specific social segments in Brazil.

## Introduction

During the early 20th century, the medical field supported an optimistic discourse on technological advances in the area and the control of infectious diseases, which for many centuries were the main causes of mortality in the world. However, several pathogens that have evolved or increased their ability to infect human beings have shown that curing communicable diseases must be a challenge for scientists for a long time to come. Examples of this are the SARS-CoV, MERS-CoV and now SARS-CoV-2 viruses, initially discovered as zoonoses which managed to change their transmission mechanism and start promoting direct human-to-human infection [1].

The infection caused by SARS-CoV-2 was classified as a differentiated clinical condition, which led to its inclusion as a new one, called Coronavirus Disease 2019 (COVID-19). The first cases were reported in the Wuhan peninsula in China, in December 2019, characterized by the high transmissibility power and cause of a severe acute respiratory dysfunction, with variations between mild, moderate and severe forms, a mortality rate growing according to older ages and to the presence of previous comorbidities, especially those that lead to some degree of cardiovascular impairment [2–3].

As of May 16, 2020, the World Health Organization has appointed 4,530,000 people infected with a total of 307,000 deaths worldwide. In Brazil, there are 220 thousand confirmed cases and 14,962 deaths caused by the disease on the same date. Which places the largest country in Latin America as the second in the world in terms of the rising epidemic curve of active cases and deaths [4].

The great challenge for Brazilian health authorities is to reduce the number of these cases to the minimum, especially the most serious ones, which require hospitalization with the support of invasive mechanical ventilation. The international experiences of countries affected by the COVID-19 epidemic demonstrate that, once the installed capacity of health services is overcome, the disease can raise its lethality rate to worrying levels, possibly leading to the death of millions of people due to the total lack health care, especially in a scenario of economic crisis and intense social inequality, which makes millions of Brazilians living in underprivileged communities more susceptible to death [5].

In Brazil, hospital bed occupancy rates are already at 100% in some states, health professionals are beginning to have to adopt patient admission criteria based on their chances of survival. Complicated decisions like this, which can mean life and death, need maximum scientific support in order to minimize the harm to all patients. Given this scenario, the present study aims to analyze the survival of inpatients and estimate the prognostic factors of patients admitted to Brazilian hospitals due to the COVID-19. Studies with this purpose, with a significant sample and with the level of scientific evidence of this design have not yet been published in the country.

Therefore, its findings may support the creation of scientifically supported clinical management protocols, minimizing conflicting situations and subsidizing decisions made in the hospital setting. In the face of a new disease that has no recognized treatment, a vaccine nor even its pathophysiology fully clarified, survival data and predictive factors are important instruments for the rational control of the epidemic and for scientific purposes, in order to better understand variables not yet discussed about COVID-19.

## Methods

### Study population and data collection

A retrospective, multicenter cohort study, of the survival analysis type, was carried out in order to estimate the prognostic factors of patients admitted to Brazilian hospitals due to COVID-19. Secondary, public and anonymous data were used from the notification forms of hospitalized patients and notified by SARS-COV-2 in hospitals in Brazil, in the observation period from February 20, 2020 to Jun 02, 2020. All hospitalized subjects and confirmed with COVID-19 through the RT-PCR exam were included in the study.

### Participants and variables

80125 hospitalizations due to COVID-19 were registered in the public national epidemiological surveillance system, obtained from the open access database [6], public and anonymous. The counting of the follow-up time starts from the first day of hospitalization until the date of death or discharge. Records of subjects with missing hospitalization dates or inconsistencies in their diagnostic record were excluded from the study. These represent 2.94% (2363/80125) of the records. Censorship occurred in patients who were discharged or transferred, however, all records used in this study had an outcome definition until Jun 02, 2020, excluding from the cohort patients who were still hospitalized, whose representation is of 39,28% of the records (19156/80125) (Figure 1).

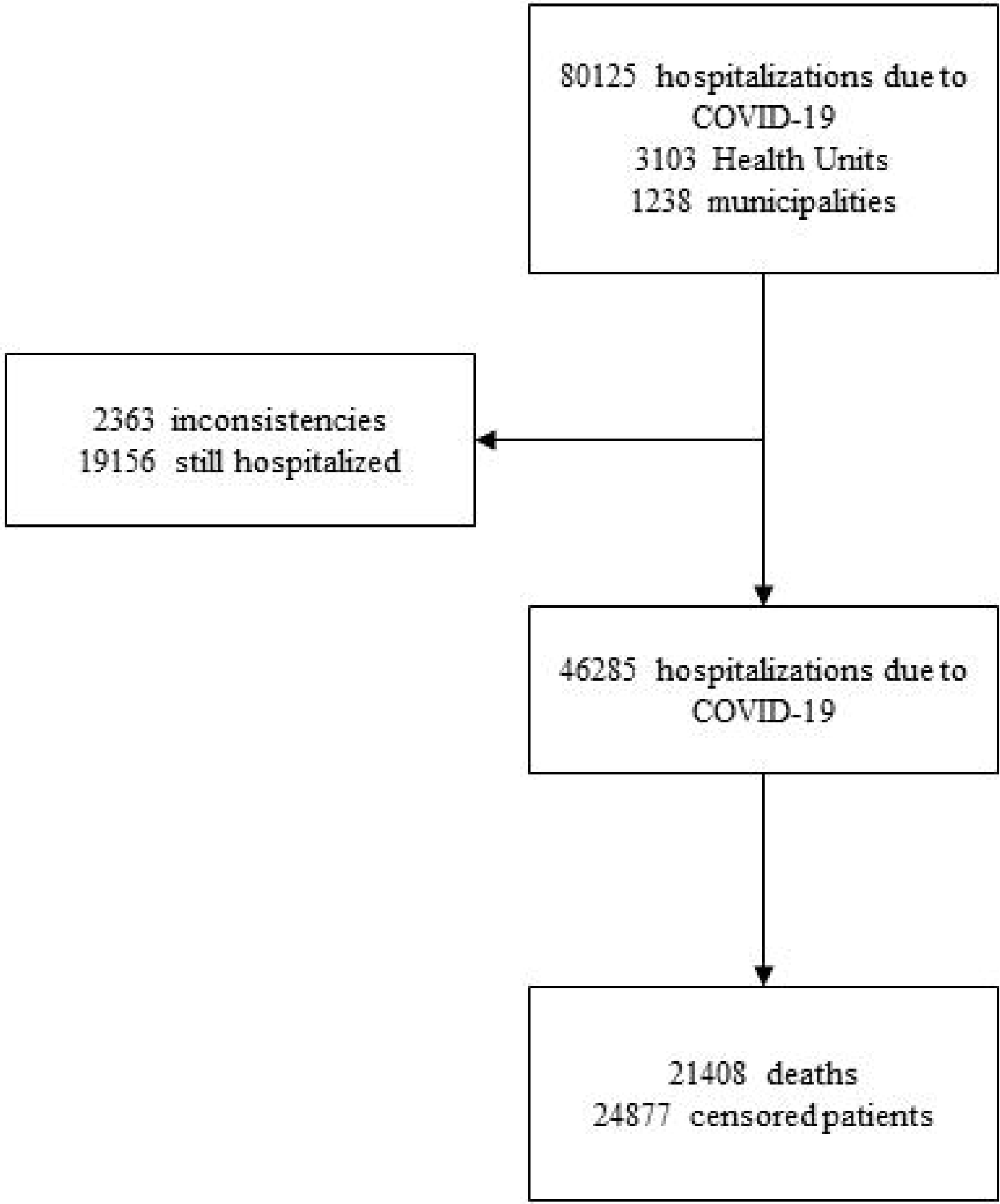

The co-variables used to compare survival curves were socioeconomic factors (age, sex, race, education and area of residence), clinical signs and symptoms (fever, cough, sore throat, diarrhea and vomiting), hospital variables (Influenza-like outbreak, hospital-acquired infection, dyspnea, respiratory distress, O2 saturation <95%, ICU admission, ICU length of stay, X-ray result), chronic disease (heart disease, hematology, down syndrome, liver disease, asthma, diabetes mellitus, neurological disease, pneumopathy, immunodepression, kidney disease, BMI), if the patient has had a flu vaccine, use of antiviral against influenza and the type of antiviral. The other variables on the notification form, whose completion was irregular or less than 5% of the cases, were excluded from the analysis (Supplementary form).

### Statistical analysis

The data were analyzed from the estimates of survival functions, using the non-parametric Kaplan-Meier method [7]. The Log-rank test and the Wilcoxon test were used to compare the survival functions for each sociodemographic, signs or symptoms, hospital and clinical covariate. The numerical variable age was categorized by tertile and the categorical variables race and education were recategorized according to the similarity of the survival curves of each category, aggregating them by theoretical category. Due to the sample size, a value of p≤0.01 was considered to accept the significant differences in the survival curves.

To assess the risk factors associated with death, Hazard Ratios (HR) and 95% confidence intervals were calculated, following Cox’s proportional hazards model [8]. Covariates with a p-value <0.250 were used to create the multiple model. The modeling was initiated by the most significant variables, both at the level of statistical significance and at the theoretical level, respecting the risk proportionality test and absence of multicollinearity to be able to test the variable in the model. The final model was tested, the device and Cox-Snell residues were analyzed. The data were analyzed using the STATA 12.0 program and in all analyzes the level of significance was considered equal to 5%.

## Results

### Survival functions

Between February 20, 2020 and Jun 02, 2020, 46285 hospitalizations due to COVID-19 were followed retrospectively. The median survival time was of 12 days (95% CI 11.82-12.18). At the end of the follow-up, 21408 deaths (46.25%) and 24877 (53.75%) censored patients were recorded (Figure 1).

The overall survival rate estimate was of 79.21% (95% CI 78.82%–79.59%) in 5 days of hospitalization and 59.22% (95% CI 58.69%–59.76%) in 10 days (Figure 2). In Table 1, when comparing the survival curves from the sociodemographic covariates, there is a significantly lower estimate of survival in 10 days in cases of ages above 68 years of 45.4% (95% CI 44.6% –46.2%), black or brown race of 51.3% (95% CI 50.3% –52.2%), living in the countryside 48.8% (95% CI 45.0%–52.5%) and illiterate 37.4% (95% CI 34.0% –40.9%).

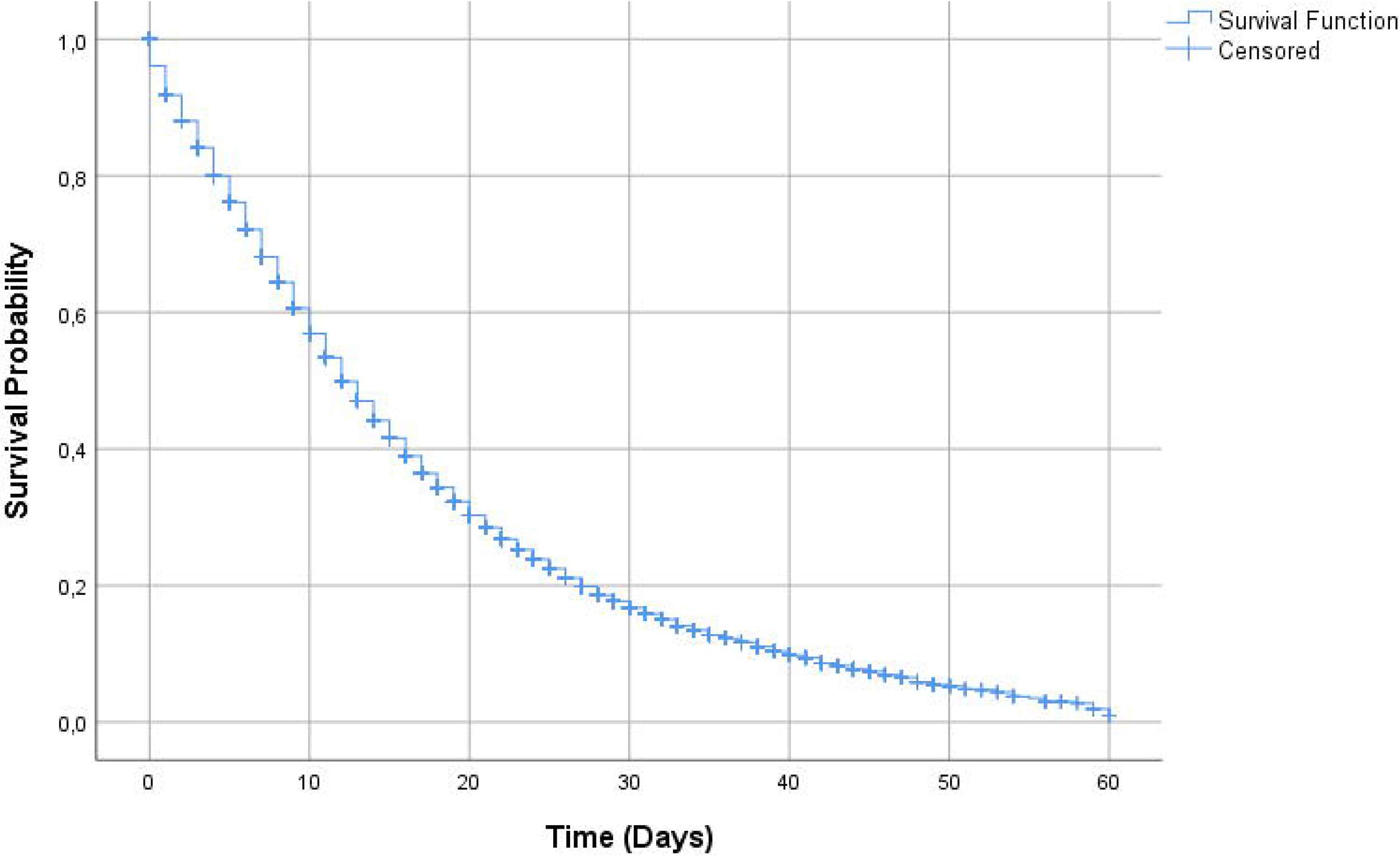

**Table 1.**
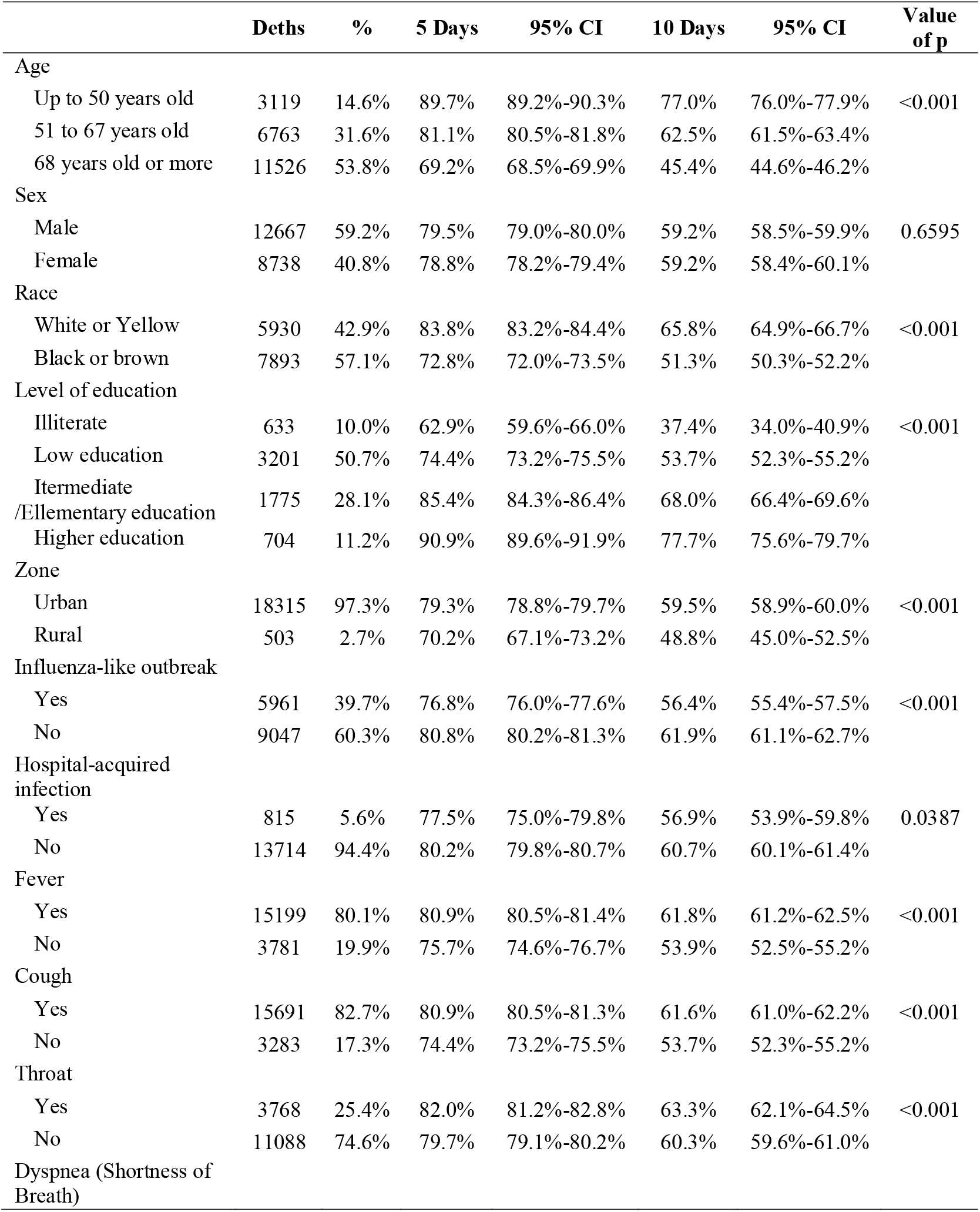
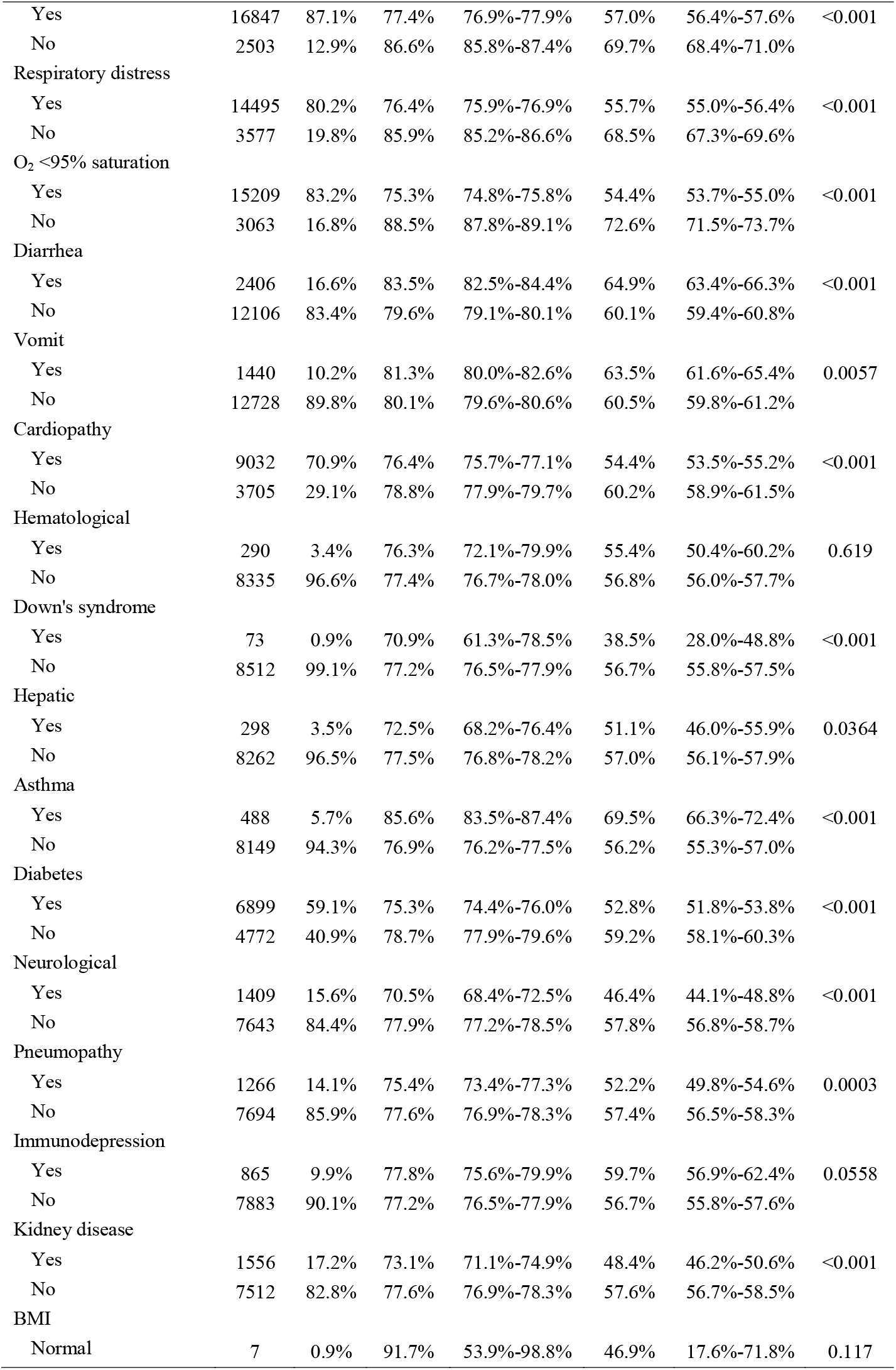
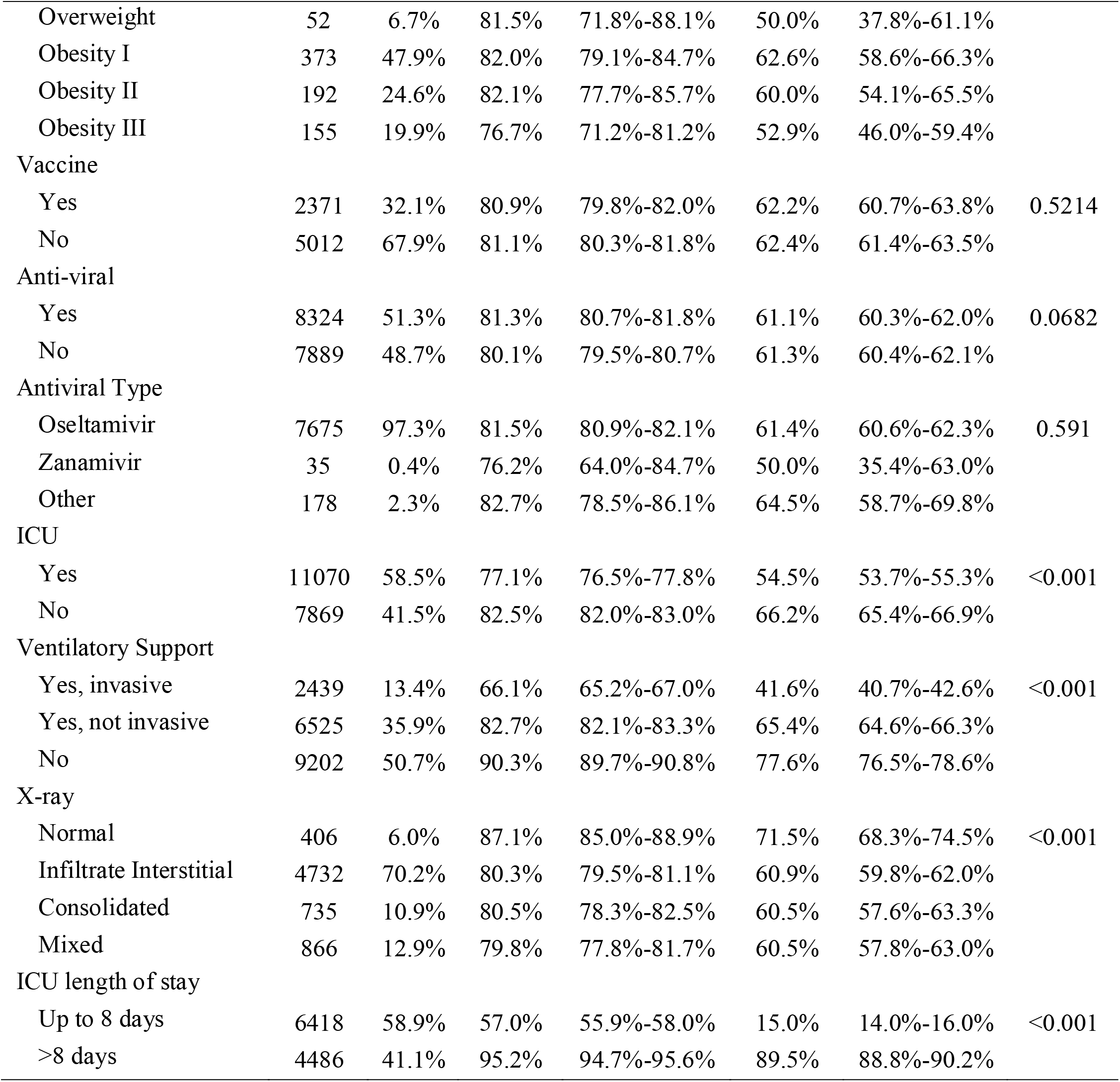
Comparison of survival estimates for patients hospitalized due to SARS-COV-2 at 5 and 10 days

Comparing at the same cutoff point in 10 days (Table 1), survival estimates were significantly lower when had dyspnea 57.0% (95% CI 56.4%–57.6%), respiratory distress 55.7% (95% CI 55.0%–56.4%), O_2_ saturation <95% 54.4% (95% CI 53.7%–55.0%), mixed X-ray alteration 60.5% (95 % CI 57.8%–63.0%), admitted to the ICU 54.5% (95% CI 53.7%–55.3%), length of permanence in the ICU up to 8 days 15.0% (95% CI 14.0%– 16.0%), invasive ventilatory support 41.6% (95% CI 40.7%–42.6%) and influenza-like outbreak 56.4% (95% CI 55.4%-57.5%). Still in Table 1, there was a lower estimate of the 10-day survival rate when the patient had chronic disease, such as heart disease 54.4% (95% CI 53.5%–55.2%), neurological 46.4% (95% CI 44.1%–48.8%), pneumopathy 52.2% (95% CI 49.8%–54.6%), diabetes 52.8% (95% CI 51.8%–53.8%) and kidney disease 48.4% (95% CI 46.2%–50.6%) and down’s syndrome 38.5% (95% CI 28.0%–48.8%).

The covariates of clinical signs showed a better estimate of the survival of patients in 10 days when they had fever 61.8% (95% CI 61.2%–62.5%), Cough 61.6% (95% CI 61.0%– 62.2%), sore throat 63.3% (95% CI 62.1%–64.5%), diarrhea 64.9% (95% CI 63.4%–66.3%) or vomiting 63.5% (95% CI 61.6%–65.4%). In the same comparison, patients who reported having asthma had a better survival estimate of 69.5% (95% CI 66.3%–72.4%).

### Cox’s proportional

In Table 2, after assessing the proportional risks, the statistically significant covariates that presented the highest risk of death were the age of 68 years HR 2.77 (95% CI 2.66–2.88), black or brown race of HR 1.54 (95% CI 1.49–1.59), living in the countryside HR 1.26 (95% CI 1.16–1.38) and illiterate HR 3.29 (95% CI 2.95–3.66), dyspnea HR 1.46 (95% CI 1.40– 1.53), respiratory distress HR 1.48 (95% CI 1.43–1.54), O_2_ saturation <95% HR 1.74 (95% CI 1.67–1.81), mixed X-ray alteration HR 1.42 (95% CI 1.26–1.59), admitted to the ICU HR 1.38 (95% CI 1.34–1.43), length of permanence in the ICU up to 8 days HR 9.15 (95% CI 8.73–9.60), invasive ventilatory support HR 2.94 (95% CI 2.81–3.07), influenza-like outbreak HR 1.18 (95% CI 1.14–1.22), heart disease HR 1.18 (95% CI 1.13–1.22), neurological HR 1.31 (95% CI 1.24–1.39), pneumopathy HR 1.11 (95% CI 1.05–1.18), diabetes HR 1.19 (95% CI 1.14–1.23), kidney disease HR 1.25 (95% CI 1.19–1.32) and down’s syndrome HR 1.51 (95% CI 1.20–1.90).

**Table 2.** Comparison of Cox proportional Hazards (HR) in relation to the risk of death

|  | **n** | **HR** | **95% CI** | **p** |
| --- | --- | --- | --- | --- |
| Age |  |  |  |  |
| Up to 50 years old | 27932 | 1.00 |  |  |
| 51 to 67 years old | 25747 | 1.74 | 1.67-1.82 | <0.001 |
| 68 years old or more | 26444 | 2.77 | 2.66-2.88 | <0.001 |
| Sex |  |  |  |  |
| Male | 45926 | 1.01 | 0.98-1.03 | 0.668 |
| Female | 34176 | 1.00 |  |  |
| Race |  |  |  |  |
| White or Yellow | 24091 | 1.00 |  |  |
| Black or brown | 26614 | 1.54 | 1.49-1.59 | <0.001 |
| Level of education |  |  |  |  |
| Higher education | 5223 | 1.00 |  |  |
| Intermediate/Elementary education | 8954 | 1.49 | 1.36-1.62 | <0.001 |
| Low education | 10391 | 2.23 | 2.06-2.42 | <0.001 |
| Illiterate | 1630 | 3.29 | 2.95-3.66 | <0.001 |
| Zone |  |  |  |  |
| Urban | 68418 | 1.00 |  |  |
| Rural | 1864 | 1.26 | 1.16-1.38 | <0.001 |
| Influenza-like outbreak |  |  |  |  |
| Yes | 19609 | 1.18 | 1.14-1.22 | <0.001 |
| No | 35911 | 1.00 |  |  |
| Hospital-acquired infection |  |  |  |  |
| Yes | 2143 | 1.07 | 1.00-1.15 | 0.044 |
| No | 52180 | 1.00 |  |  |
| Fever |  |  |  |  |
| Yes | 58977 | 0.79 | 0.77-0.83 | <0.001 |
| No | 13132 | 1.00 |  |  |
| Cough |  |  |  |  |
| Yes | 61368 | 0.79 | 0.76-0.82 | <0.001 |
| No | 10817 | 1.00 |  |  |
| Throat |  |  |  |  |
| Yes | 17084 | 0.88 | 0.85-0.91 | <0.001 |
| No | 40740 | 1.00 |  |  |
| Dyspnea (Shortness of Breath) |  |  |  |  |
| Yes | 55650 | 1.46 | 1.40-1.53 | <0.001 |
| No | 15218 | 1.00 |  |  |
| Respiratory distress |  |  |  |  |
| Yes | 46460 | 1.48 | 1.43-1.54 | <0.001 |
| No | 19326 | 1.00 |  |  |
| O<sub>2</sub> <95% saturation |  |  |  |  |
| Yes | 44351 | 1.74 | 1.67-1.81 | <0.001 |
| No | 21198 | 1.00 |  |  |
| Diarrhea |  |  |  |  |
| Yes | 10881 | 0.86 | 0.82-0.90 | <0.001 |
| No | 45191 | 1.00 |  |  |
| Vomit |  |  |  |  |
| Yes | 6128 | 0.93 | 0.88-0.98 | 0.007 |
| No | 48436 | 1.00 |  |  |
| Cardiopathy |  |  |  |  |
| Yes | 25160 | 1.18 | 1.13-1.22 | <0.001 |
| No | 12619 | 1.00 |  |  |
| Hematological |  |  |  |  |
| Yes | 759 | 1.03 | 0.91-1.16 | 0.629 |
| No | 25487 | 1.00 |  |  |
| Down's syndrome |  |  |  |  |
| Yes | 200 | 1.51 | 1.20-1.90 | <0.001 |
| No | 25911 | 1.00 |  |  |
| Hepatic |  |  |  |  |
| Yes | 680 | 1.13 | 1.01-1.26 | 0.042 |
| No | 25372 | 1.00 |  |  |
| Asthma |  |  |  |  |
| Yes | 2238 | 0.66 | 0.61-0.73 | <0.001 |
| No | 24492 | 1.00 |  |  |
| Diabetes |  |  |  |  |
| Yes | 18872 | 1.19 | 1.14-1.23 | <0.001 |
| No | 15655 | 1.00 |  |  |
| Neurological |  |  |  |  |
| Yes | 3007 | 1.31 | 1.24-1.39 | <0.001 |
| No | 24212 | 1.00 |  |  |
| Pneumopathy |  |  |  |  |
| Yes | 2935 | 1.11 | 1.05-1.18 | <0.001 |
| No | 24148 | 1.00 |  |  |
| Immunodepression |  |  |  |  |
| Yes | 2405 | 0.93 | 0.87-1.00 | 0.063 |
| No | 24237 | 1.00 |  |  |
| Kidney disease |  |  |  |  |
| Yes | 3258 | 1.25 | 1.19-1.32 | <0.001 |
| No | 23729 | 1.00 |  |  |
| BMI |  |  |  |  |
| Normal | 17 | 1.00 |  |  |
| Overweight | 139 | 1.18 | 0.53-2.61 | 0.675 |
| Obesity I | 1227 | 1.00 | 0.47-2.12 | 0.990 |
| Obesity II | 616 | 1.16 | 0.54-2.45 | 0.706 |
| Obesity III | 475 | 1.27 | 0.59-2.70 | 0.539 |
| Vaccine |  |  |  |  |
| Yes | 9914 | 1.01 | 0.97-1.07 | 0.532 |
| No | 21723 | 1.00 |  |  |
| Anti-viral |  |  |  |  |
| Yes | 26461 | 0.97 | 0.94-1.00 | 0.076 |
| No | 34881 | 1.00 |  |  |
| Antiviral Type |  |  |  |  |
| Oseltamivir | 23906 | 1.00 |  |  |
| Zanamivir | 114 | 1.13 | 0.81-1.57 | 0.473 |
| Other | 910 | 0.95 | 0.82-1.10 | 0.491 |
| ICU |  |  |  |  |
| Yes | 24284 | 1.38 | 1.34-1.43 | <0.001 |
| No | 40834 | 1.00 |  |  |
| Ventilatory Support |  |  |  |  |
| Yes, invasive | 23638 | 2.94 | 2.81-3.07 | <0.001 |
| Yes, not invasive | 27900 | 1.59 | 1.52-1.66 | <0.001 |
| No | 14806 | 1.00 |  |  |
| X-ray |  |  |  |  |
| Normal | 2173 | 1.00 |  |  |
| Infiltrate Interstitial | 16003 | 1.42 | 1.28-1.57 | <0.001 |
| Consolidated | 2277 | 1.42 | 1.26-1.61 | <0.001 |
| Mixed | 2723 | 1.42 | 1.26-1.59 | <0.001 |
| ICU length of stay |  |  |  |  |
| Up to 8 days | 9452 | 9.15 | 8.73-9.60 | <0.001 |
| >8 days | 7588 | 1.00 |  |  |

Patients had a better prognosis when they had signs or symptoms such as fever HR 0.79 (95% CI 0.77–0.83), cough HR 0.79 (95% CI 0.76–0.82), sore throat HR 0.88 (95% CI 0.85– 0.91), diarrhea HR 0.86 (95% CI 0.82–0.90), vomiting HR 0.93 (95% CI 0.88–0.98) or asthma HR 0.66 (95% CI 0.61–0.73) (Table 2). No significant difference was found in the survival curves of the sex co-variables, hospital-acquired infection, if the patient had previous hematological disease, liver disease, immunodepression, BMI, if they had been vaccinated against influenza, use antiviral or some type of antiviral.

After adjusting for confounding variables, they remained significantly in the multiple model for increased risk of death when they were admitted to the ICU HR 1.28 (95% CI 1.21–1.35), diabetes HR 1.17 (95% CI 1.11–1.24), neurological disease HR 1.34 (95% CI 1.22–1.46), kidney disease HR 1.11 (95% CI 1.02–1.21), heart disease HR 1.14 (95% CI 1.08–1.20), black or brown race of HR 1.50 (95% CI 1.43–1.58), asthma HR 0.71 (95% CI 0.61–0.81) and pneumopathy HR 1.12 (95% CI 1.02–1.23) (Figure 3).

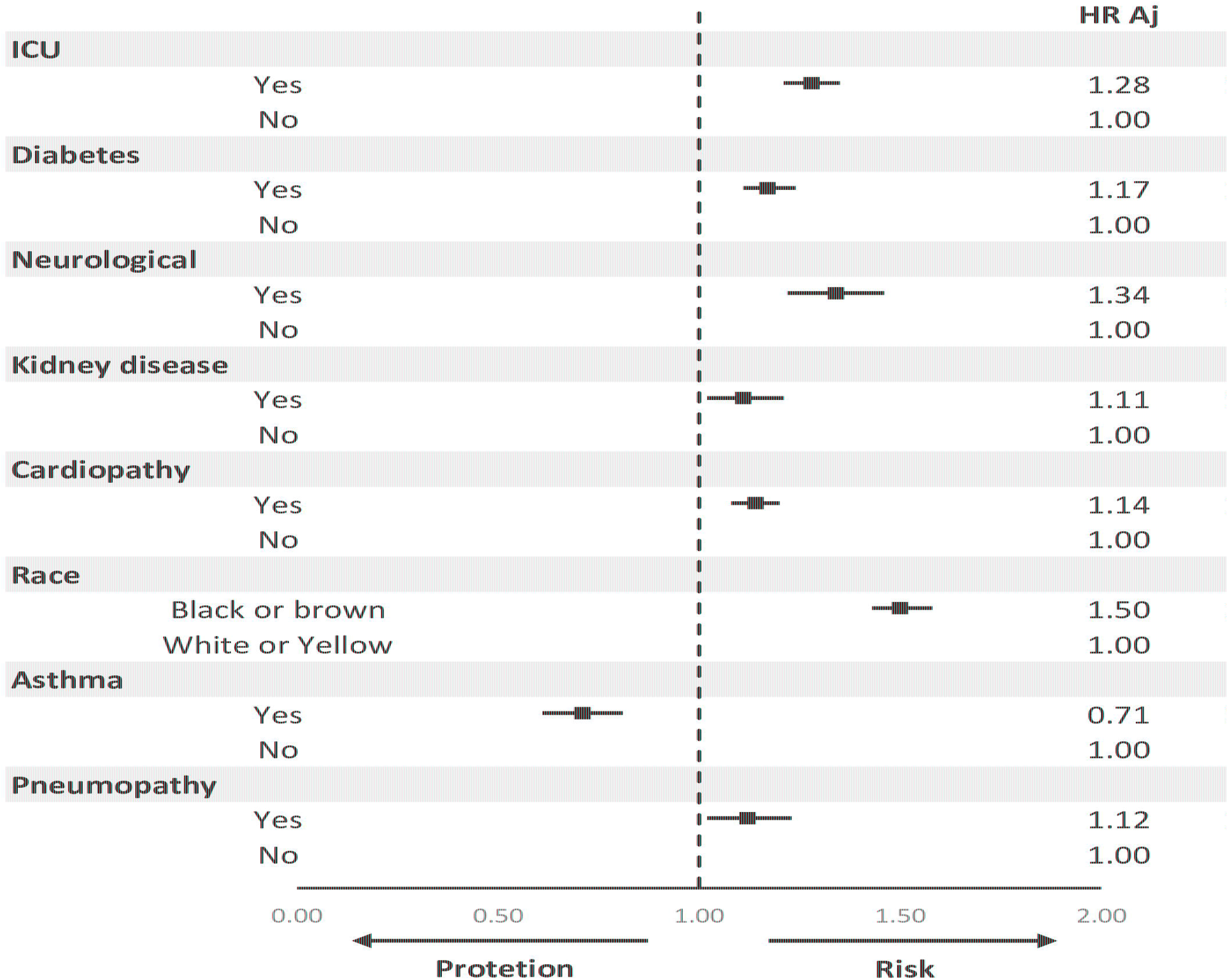

## Discussion

This multicenter retrospective cohort study of patients hospitalized with COVID-19 found important differences in survival times, as well as risk factors or protection for the death of patients in Brazilian hospitals. There was a reduced overall survival time, in addition to an association with lower survival estimates and a higher risk of death related to age, race, education, zone, dyspnea, respiratory distress, influenza-like outbreak, O_2_ saturation, X-ray changes of chest, ICU stay, length of stay in ICU, use of ventilatory support, presence of heart disease, diabetes, down’s syndrome, pneumopathy, kidney disease and neurological disease. The variables of clinical signs or symptoms, such as fever, cough, sore throat, diarrhea or vomiting, showed greater survival and less risk of death. Finally, the presence of asthma had a better prognosis.

The overall survival of patients hospitalized for treatment of COVID-19 in Brazil was 62.05%. This survival is shown to be lower than other survival studies of hospitalized patients in Brazil. A retrospective cohort study developed by Bonfada at al., whose objective was to analyze the survival of elderly patients admitted to the ICU in a low-development region, found an overall survival of 66.64%, which reinforces the concern about the lethality of COVID-19 in countries with limited resources [9].

The lack of care and clinical management protocols for patients hospitalized by COVID-19 in Brazil, closely linked to internal political disputes in the country, and the consequent delay in training the health team have a direct influence on this context. In addition, there is no strategy for testing the asymptomatic population and even for those with suggestive symptoms there is a delay in the diagnosis of about 10 days for the confirmatory result via RT-PCR, a time interval in which there is already a drop in survival for 62.05%.

In our study when comparing survivals by sociodemographic covariates, a worse prognosis was found for patients over 68 years of age, with 2.77 times the risk of death. Besides that, it was found that people with black or brown skin color had a risk of death 1.54 times bigger and people with illiterate 3.29 times. All consecutive patients diagnosed with COVID-19 admitted to the Renmin Hospital of the Wuhan University, were enrolled in a retrospective cohort study. One third of the patients improved in hospital during follow-up. Twenty-five patients died, a mortality rate of 3.77%. In this study, older patients were prone to have severe COVID-19 symptoms and unimprovement, and were more likely to die in the hospital [10]. Another cohort studies identified several risk factors for death in adults who were hospitalized with COVID-19. Older age was also associated with higher odds of in-hospital death, it revealed age IZ65 years as a strong predictor for death from COVID-19 pneumonia [11–12].

According to the Brazilian Institute of Geography and Statistics (IBGE), Brazil has a population of about 211 million inhabitants, among which there are 25 million elderly people and 55 million people who are below the poverty line, a group that is mostly composed by black people with low education [13]. Populations that live under vulnerability or social inequity often have less access to information and health services, and may reach hospital services late. In the same way, most of the people working as self-employed or underemployed, are forced out of social isolation, often without protection, to ensure the livelihood of their families, what makes them subject to potential infection rates even higher.

In this sense, health policies need to have access to the information considered above to promote an adequate confrontation with reality, especially by creating protection mechanisms for these vulnerable groups. Otherwise, the COVID-19 epidemic in Brazil runs the risk of being a catalyst that assumes a higher risk of death of 26% of its entire population, indirectly characterized as a means of eugenics, based on socioeconomic and demographic criteria, worrying and incongruent with the constitutional principles and the guarantee of the principle of equity in health care, one of the pillars of the Brazilian public health system. Underprivileged people are at higher risk of infection and death from COVID-19, and they have less access to care due to systems that treat health as a commodity and not a human right [14–15].

When assessing the context of clinical variables, significance was found for better survival when patients presented signs or symptoms at the time of hospital admission, such as fever, cough, vomiting, sore throat or diarrhea, showing they may still be in the viral replication phase, therefore, the presence of signs or symptoms in this phase could be less severe. In addition, it is likely that patients will have a less severe reaction by the immune system, which would differ from other patients who may progress abruptly to a clinical severity, without having any of these symptoms at the beginning of hospitalization.

COVID-19 pneumonia is a multistate disease with clinically relevant intermediate endpoints like severity onset. Most survival data analyses set the onset as the primary endpoint, and censor recovery or hospital discharge. However, when competing risks of severity onset are present, this analytical method induces bias. The risk of severe progression assessed without considering the competition would be overestimated because the patients who would never progress (those who discharged from hospital without progression) were treated as if they could progress [16].

It has been shown that the ICU length of stay of up to three days has the worst prognosis, which reinforces our hypothesis that patients worsen their clinical condition abruptly during hospitalization, whose admission to the ICU would have little therapeutic capacity in these more severe cases of COVID-19. However, it would be necessary to develop prospective cohort studies to monitor, mainly, the changes in the clinical signs of patients with COVID-19 and their impact on the survival of hospitalized patients and how this interferes with the length of stay in the ICU.

The level and duration of infectious virus replication are important factors in assessing the risk of transmission and guiding decisions regarding isolation of patients. Because the coronavirus RNA detection is more sensitive than the virus isolation, most studies have used qualitative or quantitative viral RNA tests as a potential marker for infectious coronavirus. The detectable SARS-CoV-2 RNA persisted for a median of 20 days in survivors and it was sustained until death in no survivors [11].

In addition to clinical symptoms, an important result found in this study was asthma as a protective factor against death. Interestingly, the prevalence of asthma in patients with COVID-19 was 0.9% in ambispective cohort study of consecutive hospitalized patient. Thus, it was speculated that TH2 immune response in patients with asthma may counter the inflammation process induced by SARS-CoV-2 infection. Further studies are required to characterize the immune response and inflammation features of COVID-19 [17].

Hospitalized patients in the No. 7 Hospital of Wuhan, clinically diagnosed as “viral pneumonia” based on their clinical symptoms (fever or respiratory symptoms) with typical changes in chest radiology, were preliminarily involved in a study. No asthmatic patient was identified in this report, and only a few patients had self□reported drug hypersensitivity and urticaria. Taken together, virus infections have been associated with acute exacerbation of asthma, and allergy may not be a risk factor for virus infection. It is plausible that this concept may also apply to SARS□CoV□2; however, having no asthma patients and no respiratory allergies does not support this concept at least in this series of 140 patients [18].

Regardless of the sociodemographic variables and signs and symptoms, when analyzing the risks in the multivariate model, patients who were admitted to the ICU or who had heart, kidney, diabetes and neurological disease as comorbidities had the worst prognosis.

Most severe patients with COVID-19 admitted to the Tongji Hospital that were retrospectively enrolled and followed-up showed rapid progression and multiple organ dysfunction. During hospitalization, a substantial proportion of patients presented cardiac injury, liver and kidney dysfunction, and hyperglycemia. This present study also revealed that hyperglycemia was related with increased mortality in patients with COVID-19. The prevalence of hyperglycemia may be associated with underlying diabetes and corticosteroid therapy [17].

Led by the National Health Commission of the People’s Republic of China, a retrospective cohort, to study COVID-19 admitted cases from 575 hospitals throughout China, was established [19]. Nonsurvivors present with a higher proportion of various coexisting chronic illness in univariate analysis. Coronary heart disease and cerebrovascular disease are confirmed to be independent risk factors for death. Another study in Wuhan Pulmonary Hospital revealed that underlying cardiovascular or cerebrovascular diseases contributed to high mortality from COVID-19 pneumonia [12].

220 patients with confirmed COVID-19 were recruited from the Union Hospital of Huazhong University of Science and Technology. The present study showed that 161 COVID-19 patients (80.5 %) were with at least one of the comorbidities, including diabetes, hypertension, hepatic disease, cardiac disease, chronic pulmonary disease and others. Interestingly, COVID-19 patients with chronic pulmonary disease, mainly Chronic obstructive pulmonary disease (COPD), obviously presented elevated risk of death. It was found that there was a trend for hypertension and diabetes to elevate the risk of death from COVID-19 pneumonia [20].

In this aspect, it is necessary to develop strategies that aim to make a consistent clinical assessment on the organic impacts of patients with SARS-VOC-2 infection and comorbidities. Because of these, of course, there is already a higher risk of death on hospitalization due to factors such as a greater inflammatory response, propensity to chronic endothelial lesions and intravascular coagulation, acid/basic blood imbalance and impaired immune response.

Although the work with secondary data notification forms has its limitations, this problem is partially solved, mainly due to the country’s urgency to maintain more accurate data in the monitoring of COVID-19, so that health institutions are able to plan actions to control the epidemic. In some cases, there was incomplete documentation of the history and symptoms in the electronic database, even after making efforts regarding feedback and recollection. Some diagnoses of co-existing illnesses were originated in self-reports from patients at their admission, which might lead to recall bias.

However, this study has a robust methodology, with appropriate data analysis and the first research in the country with all hospitalization in this gap of time. Furthermore, this research confirmed the importance of age and the presence of chronic diseases not only in the incidence of more serious cases of COVID-19, but also its effects on the drop in the survival curve of patients in hospital in Brazil. Nowadays this country is considered one of the worldwide epicenters to COVID-19.

In conclusion, it is considered important that scientifically supported protocols are developed for the management of appropriate clinical care for patients in hospital due to the COVID-19, especially in the face of a scenario where there is no consensus on pharmacological treatment or even therapeutic management. It is worth mentioning the importance in the development of new studies, such as controlled and randomized clinical trials, in order to reaffirm the results of the present study, especially regarding the variable asthma as a protective factor against death.

## Data Availability

Supplied upon request

## References

1. G. L. Gilbert, SARS, MERS and COVID-19—new threats; old lessons. International Journal of Epidemiology, (2020) 1–3. https://doi.org/10.1093/ije/dyaa061.

2. M. Xie & Q. Chen, Insight into 2019 novel coronavirus — An updated interim review and lessons from SARS-CoV and MERS-CoV. International Journal of Infectious Diseases, 94 (2020) 119–124. https://doi.org/10.1016/j.ijid.2020.03.071.

3. S. A. Meo, A. M. Alhowikan, T. A. L. Khlaiwi, I. M. Meo, D. M. Halepoto, M. Iqbal, A. M. Usmani, W. Hajjar, & N. Ahmed, Novel coronavirus 2019-nCoV: Prevalence, biological and clinical characteristics comparison with SARS-CoV and MERS-CoV. European Review for Medical and Pharmacological Sciences, 24 (2020) 2012–2019. https://doi.org/10.26355/eurrev_202002_20379.

4. WHO, Coronavirus disease. World Health Organization, 2019 (2020) 2633. https://doi.org/10.1001/jama.2020.2633.

5. The Lancet, COVID-19 in Brazil: “So what?” The Lancet, 395 (2020) 1461. https://doi.org/10.1016/S0140-6736(20)31095-3.

6. M. da S. Brasil, Open Date SARS COV 19. (2020). https://opendata.saude.gov.br/ (accessed May 10, 2020).

7. E. L. Kaplan & P. Meier, Nonparametric estimation from incomplete samples. J. of the ASA, 73 (1958) 457–481.

8. V. S. Stel, F. W. Dekker, G. Tripepi, C. Zoccali, & K. J. Jager, Survival Analysis II: Cox Regression. Nephron Clinical Practice, 119 (2011) c255–c260. https://doi.org/10.1159/000328916.

9. D. Bonfada, M. M. dos Santos, K. C. Lima, & A. Garcia-Altés, Survival analysis of elderly patients in Intensive Care Units. Revista Brasileira de Geriatria e Gerontologia, 20 (2017) 197–205. https://doi.org/10.1590/1981-22562017020.160131.

10. J. Zhang, X. Wang, X. Jia, J. Li, K. Hu, G. Chen, J. Wei, Z. Gong, C. Zhou, H. Yu, M. Yu, H. Lei, F. Cheng, B. Zhang, Y. Xu, G. Wang, & W. Dong, Risk factors for disease severity, unimprovement, and mortality in COVID-19 patients in Wuhan, China. Clinical Microbiology and Infection, (2020). https://doi.org/10.1016/j.cmi.2020.04.012.

11. F. Zhou, T. Yu, R. Du, G. Fan, Y. Liu, Z. Liu, J. Xiang, Y. Wang, B. Song, X. Gu, L. Guan, Y. Wei, H. Li, X. Wu, J. Xu, S. Tu, Y. Zhang, H. Chen, & B. Cao, Clinical course and risk factors for mortality of adult inpatients with COVID-19 in Wuhan, China: a retrospective cohort study. The Lancet, 395 (2020) 1054–1062. https://doi.org/10.1016/S0140-6736(20)30566-3.

12. R.-H. Du, L.-R. Liang, C.-Q. Yang, W. Wang, T.-Z. Cao, M. Li, G.-Y. Guo, J. Du, C.-L. Zheng, Q. Zhu, M. Hu, X.-Y. Li, P. Peng, & H.-Z. Shi, Predictors of mortality for patients with COVID-19 pneumonia caused by SARS-CoV-2: a prospective cohort study. European Respiratory Journal, 55 (2020) 2000524. https://doi.org/10.1183/13993003.00524-2020.

13. I. B. de G. e E. Brasil, População Nacional. (2020). https://www.ibge.gov.br/estatisticas/sociais/populacao.html (accessed May 13, 2020).

14. D. Chiriboga, J. Garay, P. Buss, R. S. Madrigal, & L. C. Rispel, Health inequity during the COVID-19 pandemic: a cry for ethical global leadership. The Lancet, 6736 (2020) 31145. https://doi.org/10.1016/S0140-6736(20)31145-4.

15. J. J. Van Bavel, K. Baicker, P. S. Boggio, V. Capraro, A. Cichocka, M. Cikara, M. J. Crockett, A. J. Crum, K. M. Douglas, J. N. Druckman, J. Drury, O. Dube, N. Ellemers, E. J. Finkel, J. H. Fowler, M. Gelfand, S. Han, S. A. Haslam, J. Jetten, S. Kitayama, D. Mobbs, L. E. Napper, D. J. Packer, G. Pennycook, E. Peters, R. E. Petty, D. G. Rand, S. D. Reicher, S. Schnall, A. Shariff, L. J. Skitka, S. S. Smith, C. R. Sunstein, N. Tabri, J. A. Tucker, S. van der Linden, P. van Lange, K. A. Weeden, M. J. A. Wohl, J. Zaki, S. R. Zion, & R. Willer, Using social and behavioural science to support COVID-19 pandemic response. Nature Human Behaviour, 4 (2020) 1–12. https://doi.org/10.1038/s41562-020-0884-z.

16. L. Zeng, J. Li, M. Liao, R. Hua, P. Huang, M. Zhang, Y. Zhang, Q. Shi, Z. Xia, X. Ning, D. Liu, J. Mo, Z. Zhou, Z. Li, Y. Fu, Y. Liao, J. Yuan, L. Wang, Q. He, L. Liu, & K. Qiao, Risk assessment of progression to severe conditions for patients with COVID-19 pneumonia: a single-center retrospective study. medRxiv, (2020) 2020.03.25.20043166. https://doi.org/10.1101/2020.03.25.20043166.

17. X. Li, S. Xu, M. Yu, K. Wang, Y. Tao, Y. Zhou, J. Shi, M. Zhou, B. Wu, Z. Yang, C. Zhang, J. Yue, Z. Zhang, H. Renz, X. Liu, J. Xie, M. Xie, & J. Zhao, Risk factors for severity and mortality in adult COVID-19 inpatients in Wuhan. Journal of Allergy and Clinical Immunology, (2020) 1–9. https://doi.org/10.1016/j.jaci.2020.04.006.

18. J. Zhang, X. Dong, Y. Cao, Y. Yuan, Y. Yang, Y. Yan, C. A. Akdis, & Y. Gao, Clinical characteristics of 140 patients infected with SARS_CoV_2 in Wuhan, China. Allergy, (2020) all.14238. https://doi.org/10.1111/all.14238.

19. W. Liang, W. Guan, R. Chen, W. Wang, J. Li, K. Xu, C. Li, Q. Ai, W. Lu, H. Liang, S. Li, & J. He, Cancer patients in SARS-CoV-2 infection: a nationwide analysis in China. The Lancet Oncology, 21 (2020) 335–337. https://doi.org/10.1016/S1470-2045(20)30096-6.

20. L. Fu, J. Fei, H.-X. Xiang, Y. Xiang, Z.-X. Tan, M.-D. Li, F.-F. Liu, H.-Y. Liu, L. Zheng, Y. Li, H. Zhao, & D.-X. Xu, Analysis of Death Risk Factors Among 200 COVID-19 Patients in Wuhan, China: A Hospital-Based Case-Cohort Study. SSRN Electronic Journal, 86 (2020). https://doi.org/10.2139/ssrn.3551430.

